# Place-Based Disparities in Treatment and Time-to-Initiation for Head and Neck Cancer

**DOI:** 10.1101/2025.08.05.25333057

**Authors:** Anders Erickson, Jason Semprini

**Affiliations:** Des Moines University

**Author notes:** **Corresponding Author:** Jason Semprini, 8025 Grand Ave, West Des Moines, IA 50266.

## Abstract

**Background:** In the US, 72,000+ adults are diagnosed with Head and Neck Cancer (HNC) annually. Despite improving outcomes overall, place-based disparities persist at the extremes of the care continuum. While existing evidence has emphasized the disparities at diagnosis, less is known about how patterns of care differ after diagnosis. To inform system-level strategies to improve care delivery in low-income communities, we aimed to evaluate place-based disparities in type of treatment received and time-to-treatment initiation for HNC patients.

**Methods:** We analyzed Surveillance, Epidemiological, and End Results (SEER) case data (2018–2022). Our first mutually exclusive set of binary outcomes related to the type of treatment received were categorized as surgery only, surgery with adjuvant radiotherapy, definitive chemoradiation, radiation or chemotherapy alone, or no treatment. Our second set of binary outcomes related to time from diagnosis to treatment initiation: 0–29, 30–59, 60–89, and 90+ days. Linear probability and multinomial regression models adjusted for tumor site, stage, sociodemographics, and geography to estimate the association between residing in a low-income county (<80k median household income) and differences in the probability of each outcome.

**Results:** Our sample included 70,468 HNC cases. We found no place-based differences for adjuvant or definitive treatment. Compared to patients in high-income counties, patients in low-income counties were 1.1%-points less likely to receive surgery only (−2.2, −0.1) and 1.0%-points more likely to receive radiation or chemotherapy alone (0.1,1.8); and 2.1%-points less likely to begin treatment within 0–29 days (−3.7, −0.4) and 1.4%-points more likely to delay treatment until 60–89 days (0.5, 2.3). These differences remained consistent across nearly all sociodemographic subgroups.

**Conclusion:** Our findings warrant implementing and evaluating system-level interventions to promote access to high-quality, timely HNC treatment in low-income communities. The extent to which place-based disparities in treatment influence disparities in survival remains unknown.

## Introduction

According to the National Cancer Institute, over 72,000 people in the United States will be diagnosed with cancers of the oral cavity, pharynx, or larynx in 2025, the major subtypes of head and neck cancer (HNC)[1,2]. More than 16,000 adults are expected to die from HNC[1,2]. Although the overall incidence of HNC in the U.S. has declined in recent decades, this trend masks substantial variation by HNC subsite [3]. Broadly speaking, as the incidence of oral cavity and laryngeal cancers have steadily decreased as a result of declining smoking rates, HPV-associated oropharyngeal cancers (p16+) have risen significantly[4–6]. Still, although incidence trends vary, a growing proportion of HNCs are diagnosed at later stages[7].

These dynamic trends in HNC incidence coincide with emerging disparities in HNC staging [8–11]. There are currently no recommended screening protocols for diagnosing HNCs at earlier stages, so we should not expect staging patterns to improve in the short term[12–16]. More importantly, while differences in stage at diagnosis may drive observed differences in downstream outcomes such as survival and mortality, stage at diagnosis most certainly influences treatment modalities[17–21]. Given the absence of effective screening protocols, health systems must continue relying on effective and timely treatment protocols to improve survival for patients with HNC.

Unfortunately, consistent with the observed disparities at the extremes of the cancer continuum (i.e., staging and survival), existing evidence has revealed disparities in HNC treatment[8–11,22,23]. Most of the evidence has focused on individual-level factors, specifically identifying that Black adults with HNC receive different types of treatment and initiate treatment later than other racial/ethnic groups[24–26]. These racial and socioeconomic differences in treatment appear to be associated with worse survival[27–32]. Yet, unlike the existing evidence on individual-level factors (such as race/ethnicity and poverty), the evidence investigating HNC treatment differences by community-level factors has been less conclusive[33–37]. For example, Semprini and Williams found mixed evidence that treatment outcomes vary by neighborhood, but only considered radiation therapy and a crude 2-month time-to-treatment measure as outcomes[36,38]. Rural-urban differences also appear to be driven by broader health system factors [33,34,36,37,39,40]. Yet, studies analyzing community-level resources leave a peculiar evidence gap: research analyzing state factors give little attention to county-level differences and research analyzing granular geographic factors ignore state-level differences[33]. This ambiguity hinders efforts to promote health equity. Quite simply, how can practitioners and policymakers implement effective health system interventions without knowing at which level of the system the intervention should be implemented?

### Objective

Despite decades of evidence highlighting individual-level (i.e., Black-White) disparities and the emerging appreciation for geographic factors (i.e., rural-urban), our understanding of how HNC patterns of care differ by community-level factors (i.e., high vs. low-income counties) is limited[58]. Recent attempting to evaluate community-level factors did not comprehensively evaluate treatment outcomes; account for tumor, individual, and state-level factors; or relied on methods whose results are rarely reported or interpreted correctly[33,36–38,40]. To address these gaps, we analyzed the latest population-based registry data to evaluate whether patients residing in low-income counties receive different treatment or initiate treatment later than matched counterparts residing in high-income counties. Our contributions can inform targeted health system approaches advancing high-quality, timely care in low-income communities.

## Methods

### Data and Measures

Data for this study came from the Surveillance, Epidemiology, and End Results (SEER) program, specifically the November 2024 SEER Research Plus Limited-Field Data (21 Registries)[41]. We identified patients diagnosed with HNC between 2018 to 2022. Cases were restricted to those of a single primary malignancy or cases where HNC was one of two primaries. Cases diagnosed at autopsy or death certificate only were excluded. Further, because Texas and Illinois registry data do not contain comprehensive staging or treatment information, those two states were excluded.

Each case was classified using ICD-O-3, and grouped into p16-positive oropharynx, p16-negative oropharynx, other pharynx, larynx, oral cavity, or salivary gland. For tumor characteristics, HNC staging was defined using the American Joint Committee on Cancer (AJCC) and the tumor-node-metastasis (TNM) staging system, 8th Edition[42]. Cases were further stratified into early (Stage I–II) disease, advanced (Stage III–IV) disease, or unstaged. SEER also included patient information. Demographic factors included patient sex, race/ethnicity, marital status, age at diagnosis, and state of residence. Metropolitan status was classified using the United States Department of Agriculture’s Rural-Urban Continuum Codes (RUC), which categorizes U.S. counties into nine groups based on their degree of urbanization, population size, and proximity to metropolitan areas[43,44]. SEER also reports whether the patient received treatment at a hospital accredited by the Commission on Cancer (CoC).

Our study had two sets of mutually exclusive outcome measures. The first set related to the type of treatment received. Treatment modalities were grouped into five categories: definitive CRT (radiation and chemo without surgery), primary surgery alone, radiation or chemotherapy alone, surgery plus adjuvant radiotherapy (RT), and no treatment or unknown treatment. The second set related to time-to-treatment initiation, reported in days from diagnosis. Each group was categorized into 0-29 days, 30-59 days, 60–89 days, or 90+ days.

Our primary independent variable of interest was a county-level variable measuring median household income. We classified all cases as residing in a low-income county if the median household income was below $80,000.

### Statistical Analysis

Our study aimed to estimate the independent association between residing in a low-income county and probability of 1) receiving specific treatment modalities and 2) initiating treatment within a specific timeframe. All analyses adjusted for individual factors (sex, race/ethnicity, age at diagnosis, whether the patient received treatment at an accredited hospital), geographic factors (state of residence, if the patient resided in a metropolitan county), and tumor characteristics (HNC site group, stage at diagnosis). To account for differences over time, including potential disruptions from the COVID-19 pandemic, each analysis included year fixed-effects[45]. As a sensitivity check, our time-to-treatment regression models include an alternative specification where we adjust for type of treatment received.

For each aim, we constructed two distinct tests. First, we estimated a linear probability regression model, separately, for each mutually exclusive outcome[46]. Second, we estimated a multinomial logistic regression model with a post-estimation, simultaneous marginal analysis procedure[47–49]. Both tests estimate the average association between residing in a low-income county and the probability of each respective outcome. For inference, all estimated standard errors were robust to heteroskedasticity and clustered at the state level (alpha = 0.05)[50].

In addition to estimating the overall associations, we replicated the linear regression analysis by sex (male, female), age (<65, 65+), race/ethnicity (non-Hispanic White, all other racial/ethnic groups combined), stage at diagnosis (early, advanced, unstaged), and HNC site group (OPC p16+, OPC p16-, other pharynx, oral cavity, larynx, salivary gland).

## Results

### Summary Statistics

Table 1 presents the summary statistics of our analytic sample, which includes 70,468 HNC cases. The most common HNC subtype was oral cavity cancers with 19,678 cases (27.92%), followed by HPV-positive oropharyngeal cancer (p16+), accounting for 18,815 cases (26.70%). Laryngeal cancers comprised 12,701 cases (18.02%), while HPV-negative oropharyngeal cancers (p16−) accounted for 7,220 cases (10.25%). Salivary gland tumors and other pharyngeal cancers made up 6,206 cases (8.81%) and 5,848 cases (8.30%), respectively. Regarding treatment modality, 21,525 patients (30.55%) received definitive CRT, 19,030 (27.01%) underwent surgery with adjuvant RT, 13,967 (19.82%) received primary surgery only, and 9,312 (13.21%) received either radiation or chemotherapy alone. A total of 6,634 patients (9.41%) did not receive any documented treatment. Supplemental Appendix S1 reports the case counts receiving specific treatment for each HNC type, overall and by stage at diagnosis. County-level income distribution was nearly even, with 34,573 patients (49.06%) living in low-income counties and 35,895 (50.94%) in higher-income areas.

**Table 1:** Summary Sample Characteristics – HNC Patients Receiving Treatment (SEER 2018-2022)

| Variable | N | (%) |
| --- | --- | --- |
| **HNC Type** |  |  |
| Larynx | 12,701 | (18.02%) |
| OPC P16+ | 18,815 | (26.70%) |
| OPC P16- | 7,220 | (10.25%) |
| Oral Cavity | 19,678 | (27.92%) |
| Other Pharynx | 5,848 | (8.30%) |
| Salivary | 6,206 | (8.81%) |
| **Treatment Group** |  |  |
| Definitive CRT | 21,525 | (30.55%) |
| Primary Surgery Only | 13,967 | (19.82%) |
| Radiation or Chemo Only | 9,312 | (13.21%) |
| Surgery Plus Adjuvant RT | 19,030 | (27.01%) |
| None | 6,634 | (9.41%) |
| **Patient Characteristics** |  |  |
| Age 65+ | 32,945 | (46.75%) |
| Male | 51,170 | (72.61%) |
| Non-Hispanic White | 49,765 | (70.62%) |
| Treated at CoC Accredited Hospital | 51,395 | (72.93%) |
| Metro County of Residence | 61,092 | (86.69%) |
| Low-Income County<br>(<80k household income) | 34,573 | (49.06%) |
| **Year of Diagnosis** |  |  |
| 2018 | 13,912 | (19.74%) |
| 2019 | 14,289 | (20.28%) |
| 2020 | 13,332 | (18.92%) |
| 2021 | 14,414 | (20.45%) |
| 2022 | 14,521 | (20.61%) |
| **State** |  |  |
| California | 21,057 | (29.88%) |
| Connecticut | 2,718 | (3.86%) |
| Georgia | 7,680 | (10.90%) |
| Hawaii | 1,110 | (1.58%) |
| Idaho | 1,209 | (1.72%) |
| Iowa | 2,697 | (3.83%) |
| Kentucky | 4,353 | (6.18%) |
| Louisiana | 3,958 | (5.62%) |
| New Jersey | 5,954 | (8.45%) |
| New Mexico | 1,303 | (1.85%) |
| New York | 13,418 | (19.04%) |
| Seattle (Puget Sound) | 3,729 | (5.29%) |
| Utah | 1,282 | (1.82%) |
| **Stage of Diagnosis** |  |  |
| Early (I-II) | 30,933 | (43.90%) |
| Late (III-IV) | 27,395 | (38.88%) |
| Unstaged | 12,140 | (17.23%) |
| N = 70,468 |  |  |

Table 1 reports the count (proportion) of the head and neck cancer case sample. Source: SEER 2018-2022. OPC = oropharynx cancer. RT = radiation therapy. CRT = chemoradiation therapy. CoC = Commission on Cancer.

Figure 1 visualizes the unadjusted, observed differences in treatment modality and time-to-treatment initiation by county-level income. For treatment received, there were significant differences between high- and low-income counties for each type of treatment (Supplemental Appendix S2). Patients in low-income counties were more likely to receive no treatment, perform surgery only, and receive definitive CRT. For time-to-initiation, patients in low-income counties were significantly more likely to initiate treatment 60-89 days and 90+ days from diagnosis, and significantly less likely to initiate treatment within 0-29 days (Supplemental Appendix S2).

**Figure 1.**
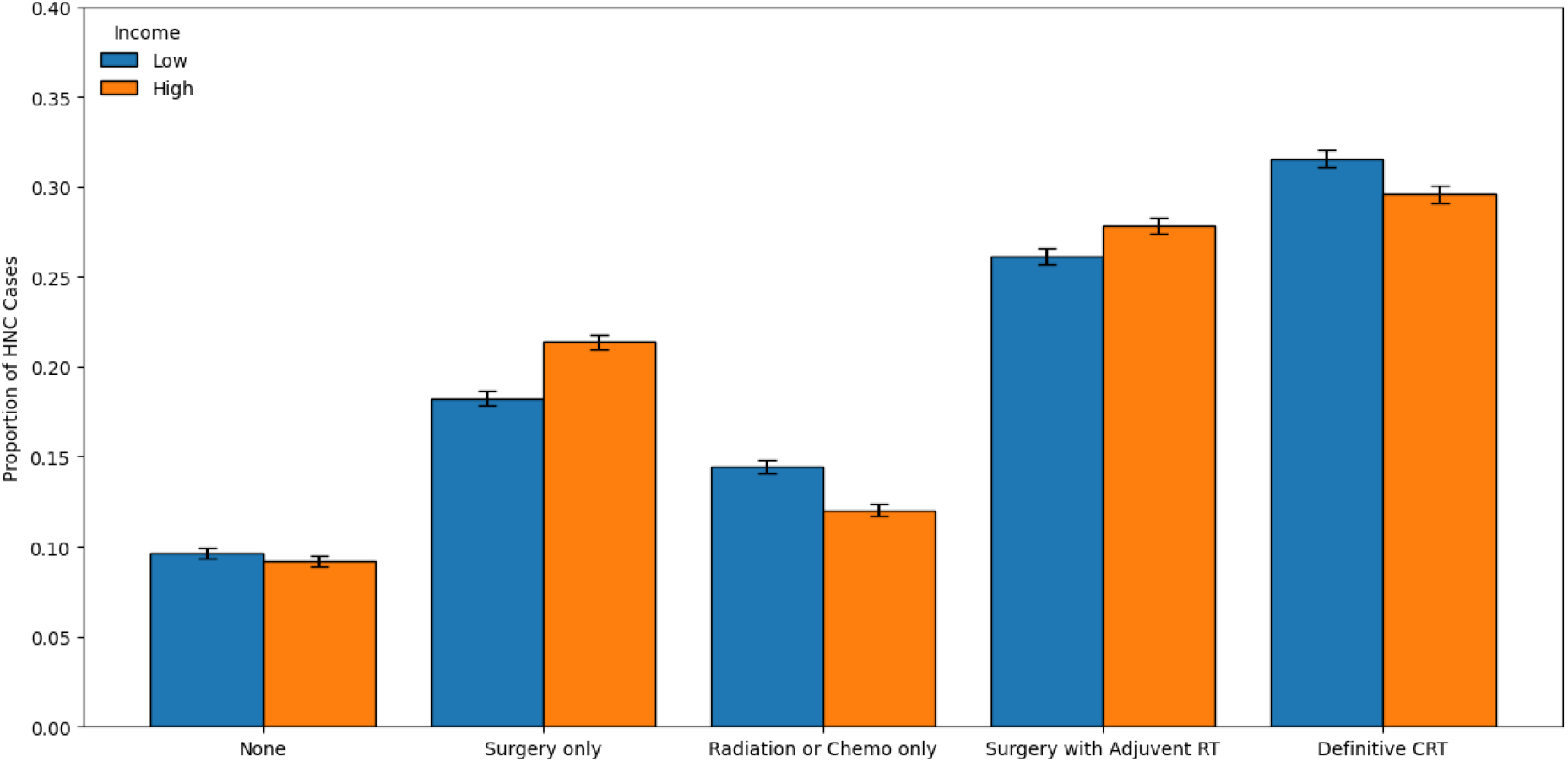
Treatment Outcomes, High and Low-Income Households. Figure 1 visualizes the proportion of the sample, by county-level median household income, receiving each type of treatment. Treatment categories are mutually exclusive. Low = Low-income county, defined as median household income less than $80k. Y-axis reported on a binary scale (0-1).

**Figure 2.**
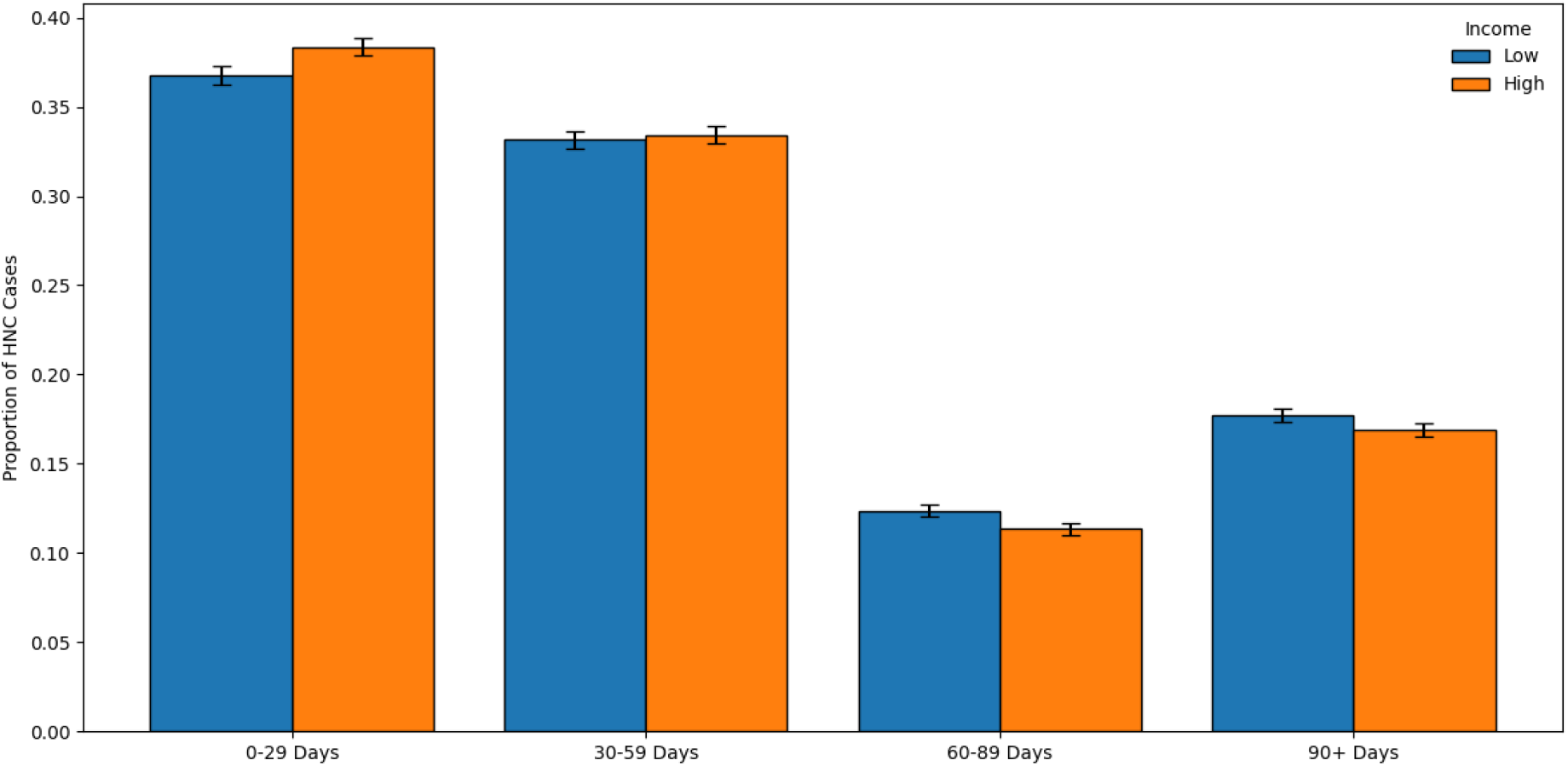
Time-to-Treatment Outcomes, High and Low-Income Households. Figure 2 visualizes the proportion of the sample, by county-level median household income, by time from diagnosis to treatment initiating. Treatment initiation categories are mutually exclusive. Low = Low-income county, defined as median household income less than $80k. Y-axis reported on a binary scale (0-1).

### Primary Results

After adjusting for tumor, individual, and geographic factors, many of the observed differences in treatment received between high and low-income counties attenuated and were no longer statistically significantly different (Table 2). For example, the estimated association between residing in a low-income county and probability of receiving no HNC treatment was statistically insignificant (Est. = -0.002, CI = -0.007, 0.003). Similarly, there was no significant association between residing in a low-income county and probability of receiving Surgery plus Adjuvant (Est. = - 0.003, CI = -0.015, 0.009) or the probability of receiving definitive CRT (Est. = 0.007, -0.006, 0.019). However, the place-based differences remained for patients receiving surgery only, as we estimated that residing in a low-income county was associated with 1.1%-point decrease in the probability of receiving surgery only (Est. = -0.011, CI = -0.022, -0.001). We also estimated that residing in a low-income county was associated with 1.0%-point increase in the probability of receiving radiation or chemo only (Est. = 0.010, CI = 0.001, 0.018).

**Table 2:** Primary Results – Association Between Residing in Low-Income County and Treatment Outcomes.

| Outcome | Outcome | LPM Estimate<br>[95% CI] | MNL Estimate<br>[95% CI] |
| --- | --- | --- | --- |
| Type of Treatment | No Treatment | -0.002<br>[-0.007, 0.003] | -0.000<br>[-0.004, 0.003] |
|  | Surgery Only | -0.011*<br>[-0.022, -0.001] | -0.011*<br>[-0.019,-0.002] |
|  | Radiation or Chemo Only | 0.010*<br>[0.001, 0.018] | 0.009*<br>[0.002, 0.016] |
|  | Surgery + Adjuvant | -0.003<br>[-0.015, 0.009] | -0.005<br>[-0.016,0.006] |
|  | Definitive CRT | 0.007<br>[-0.006, 0.019] | 0.007<br>[-0.004, 0.019] |
| Time-to-Treatment | 0-29 Days | -0.021*<br>[-0.037, -0.004] | -0.021**<br>[-0.034,-0.007] |
|  | 30-59 Days | -0.003<br>[-0.020, 0.013] | -0.004<br>[-0.018, 0.011] |
|  | 60-89 Days | 0.014**<br>[0.005, 0.023] | 0.014***<br>[0.006, 0.022] |
|  | 90+ Days | 0.010<br>[-0.013, 0.033] | 0.011<br>[-0.008, 0.030] |
| Time-to-Treatment & | 0-29 Days | -0.016<br>[-0.036,0.004] | -0.016<br>[-0.033,0.001] |
|  | 30-59 Days | -0.007<br>[-0.019,0.006] | -0.007<br>[-0.018,0.004] |
|  | 60-89 Days | 0.012*<br>[0.002,0.022] | 0.012**<br>[0.003,0.022] |
|  | 90+ Days | 0.010<br>[-0.011,0.032] | 0.010<br>[-0.007,0.028] |

Table 2 reports the primary results, estimating the association between residing in a low-income county and probability of each outcome. Outcomes are binary and estimates are reported on a binary scale, representing a percentage point change (0-1) in the probability of each outcome. LPM = Linear Probability regression model. MNL = Multinomial regression model, with postestimation marginal analysis. CI = Confidence Interval.
& Time-to-Treatment regression models adjust for type of treatment received.
\* p < 0.05, \*\* p < 0.01, \*\*\* p < 0.001.

In terms of time-to-treatment, even after adjusting for tumor, individual, and geographic factors, place-based disparities in timely treatment initiation persisted. We estimated that residing in a low-income county was associated with a 2.1%-point decline in the probability of initiating treatment within 0-29 days (Est. = -0.021, CI = -0.037, - 0.004). We also estimated that residing in a low-income county was associated with a 1.4%-point increase in the probability of initiating treatment within 60-89 days (Est. = 0.014, CI = 0.005, 0.023). Even when controlling for type of treatment received, residing in a low-income county was associated with 1.2%-point increase in the probability of delaying treatment 60-89 days (Est. = 0.012, CI = 0.002, 0.022) (Table 2).

Table 2 reports both the linear probability regression model results (preferred specification), along with the multinomial regression results. For all outcomes, there were no meaningful differences in point-estimates and our inference was consistent between both model specifications. See supplemental Appendix S3 for the Relative Risk Ratio results.

## Subgroup Results

Across nearly all subgroup categories, the statistically significant differences between high- and low-income counties were consistent (Figure 3). We found no evidence that the associations between residing in a low-income county and probability of receiving specific treatment were different between age groups, stages at diagnosis, and HNC site group. However, we did find that the association between residing in a low-income county and probability of receiving surgery only did vary by sex and race/ethnicity. While we found no statistically significant association between residing in a low-income county and probability of receiving surgery only in females (Est. = 0.003, CI = - 0.05, 0.011), we found that residing in a low-income county was associated with a 1.8%-point decline in the probability of receiving surgery only for males (Est. = -0.030, -0.016) (Supplemental Appendix S4). Further, while we found no association between residing in a low-income county and receiving surgery only for other racial/ethnic groups (Est. = -0.000, CI = -0.008, 0.008), we estimated that residing in a low-income county was associated with a 1.7%-point decrease in the probability of receiving surgery only (Est. = -0.017, CI = -0.027, -0.007) (Supplemental Appendix S4).

**Figure 3.**
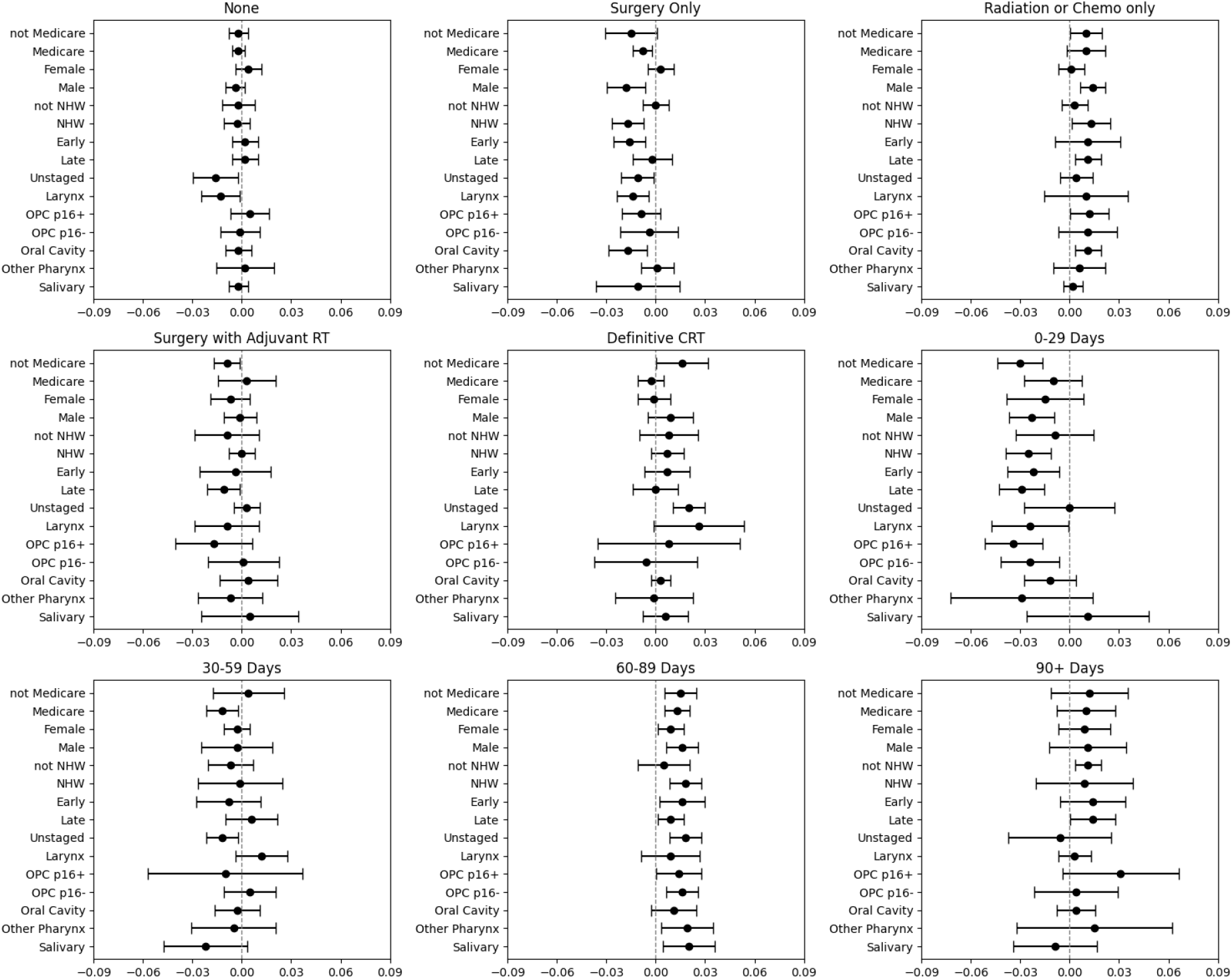
Subgroup Analyses – Association Between Residing in Low-Income County and HNC Treatment Outcomes. Figure 3 visualizes the subgroup results, estimating the association between residing in a low-income county and probability of each outcome. Outcomes are binary and estimates are reported on a binary scale, representing a percentage point change (0-1) in the probability of each outcome. Estimates derived from a Linear Probability regression model. Error bars represent 95% confidence interval of the point estimate.

For time-to-treatment initiation estimates, there were no differences across subgroups (Figure 3, Supplemental Appendix S5).

## Discussion

Our study aimed to quantify place-based disparities in HNC treatment outcomes. We first showed that while HNC patients in low-income counties receive different types of treatment than patients in high-income counties, those differences appear largely due to differences in tumor type, stage at diagnosis, and other socioeconomic factors. After adjusting for these factors, place-based disparities in the most common and highest quality HNC treatments (definitive CRT, adjuvant RT) were non-existent. This result conflicts with existing evidence (from a single state) suggesting that patients residing in higher income census tracts are more likely to receive definitive or adjuvant treatment [51]. Although, we should note that since Megwalu reports estimates as adjusted odds ratios, direct comparisons are inappropriate[51–54]. Still, future research is needed to reconcile the conflicting evidence. Future research should also examine the drivers of differences in single modality HNC treatment, and evaluate how these place-based disparities in surgery only versus radiation/chemo only contribute to downstream patient-centered outcomes.

While we found little evidence of place-based disparities in type of treatment received, our study revealed even after adjusting for tumor, stage, and other sociodemographic or geographic factors, place-based disparities in timely treatment initiation persist. In brief, despite receiving the same treatment, patients residing in low-income counties were more likely to initiate treatment 60-89 days after diagnosis than counterparts from high-income counties. Most of the existing HNC research has emphasized the importance of timely delivery of postoperative radiotherapy[55,56]. However, recent epidemiological evidence also suggests that delaying HNC treatment by two months may reduce survival[57]. Moreover, initiating treatment 60-days from diagnosis may also increase the risk of recurrence[58,59].

Recognizing the need to improve timely delivery of HNC treatment for patients in low-income communities, while maintaining appropriate quality of care, could be addressed at various levels of policymaking and at various points along the cancer care continuum. Public health professionals in low-resource counties can reduce the overall burden of HNC by promoting smoking cessation efforts. Despite overall declines in smoking rates, recent declines have been slowest or stagnate in the lowest income counties.[60]. Simply by reducing the number of future HNC cases, health systems may be able to deliver services more promptly and efficiently to fewer patients.

Once diagnosed, health systems (providers and payers) could influence timely HNC treatment delivery by expanding successful patient navigation models[61]. Although, a single model, despite proving to be effective at driving timely care or removing barriers to navigation services may work in a single setting, models in high resource communities will likely need to be tailored to fit the context in low-resource communities[62,63]. One potential model is currently being developed and evaluated in rural Appalachia[64]. The critical functions of these effective models must be disseminated broadly to promote greater diffusion of patient navigation in low-income counties.

From a policymaking perspective, one of the largest barriers to diffusion of effective programs which improve access to timely HNC care is the absence of data, causal data, quantifying the impact HNC care delays on survival, quality of life, mortality, and health expenditures. Such evidence could strengthen the advocacy efforts for evidence-based programs of service delivery models which reduce the diagnostic and treatment interval for HNC patients. Future research should also investigate the impact of delayed treatment and the use of non-surgical modalities on long-term outcomes for low-income patients, including survival rates, recurrence rates, and quality of life. Other models aside from patient navigation could enhance the coordination of care, expedite referrals, and expand access to surgical treatment, particularly in underserved settings, but evidence on such models remains limited at the population-level. In addition, large-scale prospective research is needed to understand how factors such as hospital resources, care infrastructure, and the availability of patient support services influence whether patients initiate treatment on time and complete their recommended care.

As we await greater primary and secondary prevention measures, stakeholders at various levels, from the hospital and county on up to the federal government, must advocate for and implement policies and programs that, by improving access to timely HNC care, reduce the burden of this increasingly common, heterogenous group of cancers.

### Limitations

This study is not without its limitations. First, due to the observational nature of the data, we were unable to draw causal conclusions regarding the effect of residing in a low-income county on HNC treatment outcomes. All observational analyses with secondary data are subject to this limitation. Therefore, we intentionally avoided causal language throughout the manuscript. Second, while our analysis leveraged population-based cancer registry data, we were limited by the lack of detailed treatment information. Specifically, the public-use SEER dataset does not capture the sequencing of treatment modalities, which precludes assessing access to timely guideline-concordant care. Future research should link SEER data with claims or electronic health records to explore this question further Third, our classification of low-income counties was based on an $80,000 household income threshold, which split our sample nearly in half. However, this threshold may not reflect meaningful differences in resource availability across states or regions. For example, $80,000 may represent relative affluence in some rural areas but still fall short of economic adequacy in high-cost urban settings. While we adjusted for state of residence, dynamic within-state contextual differences were not fully captured in our analysis. Fourth, our ability to conduct subgroup analyses by race and ethnicity were limited by small sample sizes, particularly for non-Hispanic Black (NHB), Hispanic, Asian American and Pacific Islander (AAPI), and American Indian and Alaska Native (AIAN) populations. As a result, we were unable to explore heterogeneity in outcomes across these groups and instead combined them into broader categories, which may mask important differences. Future research should intentionally oversample these populations and assess how place-based disparities intersect with racial and ethnic identity. Finally, although we adjusted for rurality using a metropolitan status classification, small sample sizes again limited our ability to assess rural subgroups separately. Understanding how rurality and community resources intersect to shape HNC treatment outcomes remains an important area for future inquiry. These limitations should be considered when interpreting the findings.

## Conclusions

Despite improving outcomes overall, place-based disparities persist in staging and survival for patients with head and neck cancer. By analyzing population-based data we found that HNC patients in low-income counties do receive different types of treatment than patients in high-income counties. However, much of those disparities are explained by differences in staging and tumor site. Specifically, we found no association between residing in a low-income county and receipt of surgery plus adjuvant radiation or definitive chemoradiation. However, even after adjustment, we found disparities in timely treatment for patients in low-income. Clearly, more work is needed to ensure that patients from low-income counties can receive timely treatment for this heterogenous group of cancers.

## Data Availability

Data availability limited to third-party restrictions.

