## Supplemental Files for "Place-Based Disparities in Treatment and Time-to-Initiation for Head and Neck Cancer"

### Supplemental Appendix S1: HNC Case Counts, by Treatment Group, Site, and Stage

#### i) All Stages

| | Larynx | OPC p16+ | OPC p16- | Oral Cavity | Other Pharynx | Salivary | Total |
|---|---|---|---|---|---|---|---|
| Definitive CRT | 3,405 | 10,364 | 3,399 | 803 | 3,359 | 195 | 21,525 |
| None | 1,304 | 699 | 1,026 | 2,033 | 848 | 724 | 6,634 |
| Primary Surgery Only | 1,546 | 1,106 | 580 | 8,576 | 125 | 2,034 | 13,967 |
| Radiation or Chemo | 3,678 | 2,189 | 1,168 | 905 | 1,012 | 360 | 9,312 |
| Surgery Plus Adjuvant | 2,768 | 4,457 | 1,047 | 7,361 | 504 | 2,893 | 19,030 |
| Total | 12,701 | 18,815 | 7,220 | 19,678 | 5,848 | 6,206 | 70,468 |

#### ii) Early Stages

| | Larynx | OPC p16+ | OPC p16- | Oral Cavity | Other Pharynx | Salivary | Total |
|---|---|---|---|---|---|---|---|
| Definitive CRT | 438 | 6,924 | 89 | 32 | 588 | 4 | 8,075 |
| None | 316 | 330 | 19 | 236 | 74 | 91 | 1,066 |
| Primary Surgery Only | 1,073 | 1,007 | 176 | 6,165 | 23 | 1,452 | 9,896 |
| Radiation or Chemo | 2,633 | 1,399 | 141 | 132 | 204 | 38 | 4,547 |
| Surgery Plus Adjuvant | 1,088 | 3,785 | 107 | 1,212 | 84 | 1,073 | 7,349 |
| Total | 5,548 | 13,445 | 532 | 7,777 | 973 | 2,658 | 30,933 |

#### iii) Advanced Stages

| | Larynx | OPC p16+ | OPC p16- | Oral Cavity | Other Pharynx | Salivary | Total |
|---|---|---|---|---|---|---|---|
| Definitive CRT | 2,717 | 1,716 | 2,413 | 700 | 2,192 | 173 | 9,911 |
| None | 467 | 153 | 418 | 574 | 284 | 210 | 2,106 |
| Primary Surgery Only | 279 | 19 | 94 | 1,224 | 28 | 393 | 2,037 |
| Radiation or Chemo .. | 765 | 422 | 618 | 646 | 512 | 265 | 3,228 |
| Surgery Plus Adjuvant | 1,506 | 211 | 515 | 5,956 | 251 | 1,674 | 10,113 |
| Total | 5,734 | 2,521 | 4,058 | 9,100 | 3,267 | 2,715 | 27,395 |

#### iv) Unstaged

| | Larynx | OPC p16+ | OPC p16- | Oral Cavity | Other Pharynx | Salivary | Total |
|---|---|---|---|---|---|---|---|
| Definitive CRT | 250 | 1,724 | 897 | 71 | 579 | 18 | 3,539 |
| None | 521 | 216 | 589 | 1,223 | 490 | 423 | 3,462 |
| Primary Surgery Only | 194 | 80 | 310 | 1,187 | 74 | 189 | 2,034 |
| Radiation or Chemo .. | 280 | 368 | 409 | 127 | 296 | 57 | 1,537 |
| Surgery Plus Adjuvant | 174 | 461 | 425 | 193 | 169 | 146 | 1,568 |
| Total | 1,419 | 2,849 | 2,630 | 2,801 | 1,608 | 833 | 12,140 |

**Supplemental Appendix S2: Proportion of HNC Cases Receiving Treatment Type and Initiating Treatment Within Specific Time, by county-level income (for Figure 1-2).**

| Outcome | Income | Estimate | Lower | Upper | p |
|---|---|---|---|---|---|
| **Treatment Type** | | | | | |
| None | High | 0.092 | 0.089 | 0.095 | |
| | Low | 0.096 | 0.093 | 0.100 | 0.041 |
| Surgery only | High | 0.214 | 0.209 | 0.218 | |
| | Low | 0.182 | 0.178 | 0.186 | < 0.001 |
| Radiation or Chemo only | High | 0.120 | 0.117 | 0.124 | |
| | Low | 0.144 | 0.141 | 0.148 | < 0.001 |
| Surgery with Adjuvant RT | High | 0.278 | 0.274 | 0.283 | |
| | Low | 0.261 | 0.257 | 0.266 | < 0.001 |
| Definitive CRT | High | 0.296 | 0.291 | 0.301 | |
| | Low | 0.316 | 0.311 | 0.320 | < 0.001 |
| **Time-to-Treatment** | | | | | |
| 0-29 Days | High | 0.383 | 0.378 | 0.389 | |
| | Low | 0.368 | 0.363 | 0.373 | < 0.001 |
| 30-59 Days | High | 0.334 | 0.329 | 0.339 | |
| | Low | 0.331 | 0.327 | 0.336 | 0.452 |
| 60-89 Days | High | 0.113 | 0.110 | 0.117 | |
| | Low | 0.124 | 0.120 | 0.127 | < 0.001 |
| 90+ Days | High | 0.169 | 0.165 | 0.173 | |
| | Low | 0.177 | 0.173 | 0.181 | 0.004 |

Outcomes are binary and estimates are reported on a binary scale (0-1). Low-income defined as median household income < $80k. p-value indicates t-test if proportions for each outcome vary between high and low-income group.

**Supplemental Appendix S3: Relative Risk Ratio (RRR) Estimates from Multinomial Logit Regression**

| Outcome | RRR | [95% CI] | |
|---|---|---|---|
| **Treatment Type** | | | |
| No Treatment | | REFERENCE | |
| Primary Surgery Only | 0.91* | [0.85, | 0.99] |
| Radiation or Chemo Only | 1.09 | [0.99, | 1.20] |
| Surgery Plus Adjuvant | 0.98 | [0.93, | 1.03] |
| Definitive CRT | 1.06 | [0.96, | 1.16] |
| **Time-to-Treatment** | | | |
| 0-29 Days | | REFERENCE | |
| 30-59 Days | 1.05* | [1.01, | 1.09] |
| 60-89 Days | 1.19** | [1.08, | 1.32] |
| 90+ Days | 1.14 | [0.97, | 1.35] |

*p < 0.05, ** p < 0.01

**Supplemental Appendix S4: Subgroup Estimates (for figure 3) – Treatment Type Outcomes**

| Subgroup | Outcome | Est. | 95% CI | |
|---|---|---|---|---|
| not Medicare | None | -0.002 | -0.008 | 0.004 |
| Medicare | None | -0.002 | -0.006 | 0.002 |
| Female | None | 0.004 | -0.004 | 0.012 |
| Male | None | -0.004 | -0.010 | 0.002 |
| not NHW | None | -0.002 | -0.012 | 0.008 |
| NHW | None | -0.003 | -0.011 | 0.005 |
| Early | None | 0.002 | -0.006 | 0.010 |
| Late | None | 0.002 | -0.006 | 0.010 |
| Unstaged | None | -0.016* | -0.030 | -0.002 |
| Larynx | None | -0.013* | -0.025 | -0.001 |
| OPC p16+ | None | 0.005 | -0.007 | 0.017 |
| OPC p16- | None | -0.001 | -0.013 | 0.011 |
| Oral Cavity | None | -0.002 | -0.010 | 0.006 |
| Other Pharynx | None | 0.002 | -0.016 | 0.020 |
| Salivary | None | -0.002 | -0.008 | 0.004 |
| not Medicare | Surgery Only | -0.015 | -0.031 | 0.001 |
| Medicare | Surgery Only | -0.008* | -0.014 | -0.002 |
| Female | Surgery Only | 0.003 | -0.005 | 0.011 |
| Male | Surgery Only | -0.018* | -0.030 | -0.006 |
| not NHW | Surgery Only | -0.000 | -0.008 | 0.008 |
| NHW | Surgery Only | -0.017** | -0.027 | -0.007 |
| Early | Surgery Only | -0.016** | -0.026 | -0.006 |
| Late | Surgery Only | -0.002 | -0.014 | 0.010 |
| Unstaged | Surgery Only | -0.011* | -0.021 | -0.001 |
| Larynx | Surgery Only | -0.014* | -0.024 | -0.004 |
| OPC p16+ | Surgery Only | -0.009 | -0.021 | 0.003 |
| OPC p16- | Surgery Only | -0.004 | -0.022 | 0.014 |
| Oral Cavity | Surgery Only | -0.017* | -0.029 | -0.005 |
| Other Pharynx | Surgery Only | 0.001 | -0.009 | 0.011 |
| Salivary | Surgery Only | -0.011 | -0.036 | 0.014 |
| not Medicare | Radiation or Chemo only | 0.010 | 0.000 | 0.020 |
| Medicare | Radiation or Chemo only | 0.010 | -0.002 | 0.022 |
| Female | Radiation or Chemo only | 0.001 | -0.007 | 0.009 |
| Male | Radiation or Chemo only | 0.014** | 0.006 | 0.022 |
| not NHW | Radiation or Chemo only | 0.003 | -0.005 | 0.011 |
| NHW | Radiation or Chemo only | 0.013 | 0.001 | 0.025 |
| Early | Radiation or Chemo only | 0.011 | -0.009 | 0.031 |
| Late | Radiation or Chemo only | 0.011* | 0.003 | 0.019 |
| Unstaged | Radiation or Chemo only | 0.004 | -0.006 | 0.014 |
| Larynx | Radiation or Chemo only | 0.010 | -0.015 | 0.035 |

| | | | | |
|---|---|---|---|---|
| OPC p16+ | Radiation or Chemo only | 0.012 | 0.000 | 0.024 |
| OPC p16- | Radiation or Chemo only | 0.011 | -0.007 | 0.029 |
| Oral Cavity | Radiation or Chemo only | 0.011* | 0.003 | 0.019 |
| Other Pharynx | Radiation or Chemo only | 0.006 | -0.010 | 0.022 |
| Salivary | Radiation or Chemo only | 0.002 | -0.004 | 0.008 |
| not Medicare | Surgery with Adjuvant RT | -0.009* | -0.017 | -0.001 |
| Medicare | Surgery with Adjuvant RT | 0.003 | -0.015 | 0.021 |
| Female | Surgery with Adjuvant RT | -0.007 | -0.019 | 0.005 |
| Male | Surgery with Adjuvant RT | -0.001 | -0.011 | 0.009 |
| not NHW | Surgery with Adjuvant RT | -0.009 | -0.029 | 0.011 |
| NHW | Surgery with Adjuvant RT | 0.000 | -0.008 | 0.008 |
| Early | Surgery with Adjuvant RT | -0.004 | -0.026 | 0.018 |
| Late | Surgery with Adjuvant RT | -0.011* | -0.021 | -0.001 |
| Unstaged | Surgery with Adjuvant RT | 0.003 | -0.005 | 0.011 |
| Larynx | Surgery with Adjuvant RT | -0.009 | -0.029 | 0.011 |
| OPC p16+ | Surgery with Adjuvant RT | -0.017 | -0.041 | 0.007 |
| OPC p16- | Surgery with Adjuvant RT | 0.001 | -0.021 | 0.023 |
| Oral Cavity | Surgery with Adjuvant RT | 0.004 | -0.014 | 0.022 |
| Other Pharynx | Surgery with Adjuvant RT | -0.007 | -0.027 | 0.013 |
| Salivary | Surgery with Adjuvant RT | 0.005 | -0.024 | 0.034 |
| not Medicare | Definitive CRT | 0.016 | 0.000 | 0.032 |
| Medicare | Definitive CRT | -0.003 | -0.011 | 0.005 |
| Female | Definitive CRT | -0.001 | -0.011 | 0.009 |
| Male | Definitive CRT | 0.009 | -0.005 | 0.023 |
| not NHW | Definitive CRT | 0.008 | -0.010 | 0.026 |
| NHW | Definitive CRT | 0.007 | -0.003 | 0.017 |
| Early | Definitive CRT | 0.007 | -0.007 | 0.021 |
| Late | Definitive CRT | -0.000 | -0.014 | 0.014 |
| Unstaged | Definitive CRT | 0.020** | 0.010 | 0.030 |
| Larynx | Definitive CRT | 0.026 | -0.001 | 0.053 |
| OPC p16+ | Definitive CRT | 0.008 | -0.035 | 0.051 |
| OPC p16- | Definitive CRT | -0.006 | -0.037 | 0.025 |
| Oral Cavity | Definitive CRT | 0.003 | -0.003 | 0.009 |
| Other Pharynx | Definitive CRT | -0.001 | -0.025 | 0.023 |
| Salivary | Definitive CRT | 0.006 | -0.008 | 0.020 |

Reports the estimated association between residing in a low-income county and change in probability of outcome.
*$p < 0.05$, ** $p < 0.01$

**Supplemental Appendix S5: Subgroup Estimates (for figure 3) – Time-to-Treatment Outcomes**

| Subgroup | Outcome | Est. | 95% CI | |
|---|---|---|---|---|
| not Medicare | 0-29 Days | -0.030** | -0.044 | -0.016 |
| Medicare | 0-29 Days | -0.010 | -0.028 | 0.008 |
| Female | 0-29 Days | -0.015 | -0.039 | 0.009 |
| Male | 0-29 Days | -0.023** | -0.037 | -0.009 |
| not NHW | 0-29 Days | -0.009 | -0.033 | 0.015 |
| NHW | 0-29 Days | -0.025** | -0.039 | -0.011 |
| Early | 0-29 Days | -0.022* | -0.038 | -0.006 |
| Late | 0-29 Days | -0.029** | -0.043 | -0.015 |
| Unstaged | 0-29 Days | -0.000 | -0.027 | 0.027 |
| Larynx | 0-29 Days | -0.024 | -0.048 | 0.000 |
| OPC p16+ | 0-29 Days | -0.034** | -0.052 | -0.016 |
| OPC p16- | 0-29 Days | -0.024* | -0.042 | -0.006 |
| Oral Cavity | 0-29 Days | -0.012 | -0.028 | 0.004 |
| Other Pharynx | 0-29 Days | -0.029 | -0.072 | 0.014 |
| Salivary | 0-29 Days | 0.011 | -0.026 | 0.048 |
| not Medicare | 30-59 Days | 0.004 | -0.018 | 0.026 |
| Medicare | 30-59 Days | -0.012* | -0.022 | -0.002 |
| Female | 30-59 Days | -0.003 | -0.011 | 0.005 |
| Male | 30-59 Days | -0.003 | -0.025 | 0.019 |
| not NHW | 30-59 Days | -0.007 | -0.021 | 0.007 |
| NHW | 30-59 Days | -0.001 | -0.026 | 0.024 |
| Early | 30-59 Days | -0.008 | -0.028 | 0.012 |
| Late | 30-59 Days | 0.006 | -0.010 | 0.022 |
| Unstaged | 30-59 Days | -0.012* | -0.022 | -0.002 |
| Larynx | 30-59 Days | 0.012 | -0.004 | 0.028 |
| OPC p16+ | 30-59 Days | -0.010 | -0.057 | 0.037 |
| OPC p16- | 30-59 Days | 0.005 | -0.011 | 0.021 |
| Oral Cavity | 30-59 Days | -0.003 | -0.017 | 0.011 |
| Other Pharynx | 30-59 Days | -0.005 | -0.030 | 0.020 |
| Salivary | 30-59 Days | -0.022 | -0.047 | 0.003 |
| not Medicare | 60-89 Days | 0.015** | 0.005 | 0.025 |
| Medicare | 60-89 Days | 0.013* | 0.005 | 0.021 |
| Female | 60-89 Days | 0.009 | 0.001 | 0.017 |
| Male | 60-89 Days | 0.016** | 0.006 | 0.026 |
| not NHW | 60-89 Days | 0.005 | -0.011 | 0.021 |
| NHW | 60-89 Days | 0.018** | 0.008 | 0.028 |
| Early | 60-89 Days | 0.016* | 0.002 | 0.030 |
| Late | 60-89 Days | 0.009 | 0.001 | 0.017 |
| Unstaged | 60-89 Days | 0.018** | 0.008 | 0.028 |
| Larynx | 60-89 Days | 0.009 | -0.009 | 0.027 |

| | | | | |
|---|---|---|---|---|
| OPC p16+ | 60-89 Days | 0.014 | 0.000 | 0.028 |
| OPC p16- | 60-89 Days | 0.016* | 0.006 | 0.026 |
| Oral Cavity | 60-89 Days | 0.011 | -0.003 | 0.025 |
| Other Pharynx | 60-89 Days | 0.019* | 0.003 | 0.035 |
| Salivary | 60-89 Days | 0.020* | 0.004 | 0.036 |
| not Medicare | 90+ Days | 0.012 | -0.012 | 0.036 |
| Medicare | 90+ Days | 0.010 | -0.008 | 0.028 |
| Female | 90+ Days | 0.009 | -0.007 | 0.025 |
| Male | 90+ Days | 0.011 | -0.013 | 0.035 |
| not NHW | 90+ Days | 0.011* | 0.003 | 0.019 |
| NHW | 90+ Days | 0.009 | -0.020 | 0.038 |
| Early | 90+ Days | 0.014 | -0.006 | 0.034 |
| Late | 90+ Days | 0.014 | 0.000 | 0.028 |
| Unstaged | 90+ Days | -0.006 | -0.037 | 0.025 |
| Larynx | 90+ Days | 0.003 | -0.007 | 0.013 |
| OPC p16+ | 90+ Days | 0.031 | -0.004 | 0.066 |
| OPC p16- | 90+ Days | 0.004 | -0.021 | 0.029 |
| Oral Cavity | 90+ Days | 0.004 | -0.008 | 0.016 |
| Other Pharynx | 90+ Days | 0.015 | -0.032 | 0.062 |
| Salivary | 90+ Days | -0.009 | -0.034 | 0.016 |

Reports the estimated association between residing in a low-income county and change in probability of outcome.
*$p < 0.05$, ** $p < 0.01$
